# COVID-19 Global Pandemic Planning: Decontamination and Reuse Processes for N95 Respirators

**DOI:** 10.1101/2020.04.09.20060129

**Authors:** Douglas J Perkins, Steven Villescas, Terry H. Wu, Timothy Muller, Steven Bradfute, Ivy Hurwitz, Qiuying Cheng, Hannah Wilcox, Myissa Weiss, Chris Bartlett, Jens Langsjoen, Phil Seidenberg

## Abstract

Coronavirus disease 2019 (COVID-19) is an illness caused by a novel coronavirus, severe acute respiratory syndrome coronavirus 2 (SARS-CoV-2). The disease was first identified as a cluster of respiratory illness in Wuhan City, Hubei Province, China in December 2019, and has rapidly spread across the globe to greater than 200 countries. Healthcare providers are at an increased risk for contracting the disease due to occupational exposure and require appropriate personal protective equipment (PPE), including N95 respirators. The rapid worldwide spread of high numbers of COVID-19 cases has facilitated the need for a substantial supply of PPE that is largely unavailable in many settings, thereby creating critical shortages. Creative solutions for the decontamination and safe reuse of PPE to protect our frontline healthcare personnel are essential. Here, we describe the development of a process that began in late February 2020 for selecting and implementing the use of hydrogen peroxide vapor (HPV) as viable method to reprocess N95 respirators. Since pre-existing HPV decontamination chambers were not available, we optimized the sterilization process in an operating room after experiencing initial challenges in other environments. Details are provided about the prioritization and implementation of processes for collection and storage, pre-processing, HPV decontamination, and post-processing of filtering facepiece respirators (FFRs). Important lessons learned from this experience include, developing an adequate reserve of PPE for effective reprocessing and distribution, and identifying a suitable location with optimal environmental controls (i.e., operating room). Collectively, information presented here provides a framework for other institutions considering decontamination procedures for N95 respirators.

## INTRODUCTION

Rapid global dissemination of a novel coronavirus disease (COVID-19) caused by an the enveloped non-segmented positive-sense RNA virus, SARS-CoV-2, has overwhelmed healthcare systems around the world. The rapid increase in clinical cases presenting at healthcare facilities when the disease propagates in a particular geographic region requires a rapid response by the healthcare system. The primary means of protecting frontline healthcare personnel (HCP) from contracting COVID-19 is through the proper use of personal protective equipment (PPE), such as N95 filtering facepiece respirators (FFRs). Based on the rapid spread of the virus around the globe, there is a high-volume demand for the continuous supply of PPE. The consequences of such a global demand has created a significant strain on the supply-chain of N95 respirators and other PPE. The shortage of PPE raises substantial concerns for healthcare facilities and HCP. The Centers for Disease Control and Prevention (CDC) has implemented an ongoing and continually updated release of information to optimize the supply of N95 respirators with most recent updates on 4 April 2020 ^1^. While it is without question that reuse of N95 respirators (and other PPE) would be obviated if an adequate supply were available, creative strategies are required when there is an imbalance in the supply and demand. Given the current global shortage of PPE, creative solutions are immediately required to mitigate the risk of exposure of HCP to SARS-CoV-2. In anticipation of such a shortage, we began exploring the most viable and safe methods for sterilizing PPE for reuse in late February 2020 at the University of New Mexico (UNM). During this short period of time, we have quickly learned the importance of having concerted and coordinated efforts devoted to the overall workflow for the safe collection, storage, decontamination, and distribution of reprocessed PPE, along with requisite safety training of staff who perform the reprocessing.

## PROCESS, METHODS, AND MATERIALS

### Selection and Prioritization of Decontamination Methods

In preparation for a probable shortage of PPE at our study sites in Kenya, and a possible shortage in the US (including UNM), we began investigating methods for decontaminating of FFRs in late February 2020. At that time, it became apparent that several decontamination procedures had been investigated, and that some of the methods (importantly) did not substantially impact on the structural integrity (i.e., filter aerosol penetration, airflow resistance, and physical integrity) of the N95 respirators after multiple decontamination cycles. In considering the possible options, we used a data-driven approach based on the currently available peer-reviewed literature, publicly available information, and consultation with subject matter experts. The strategic planning also considered the availability of instruments commonly found in in healthcare systems that could be rapidly transitioned and implemented for decontamination of N95 respirators.

Several studies have investigated common decontamination methods: Ultraviolet Germicidal Irradiation (UVGI), Hydrogen Peroxide Vapor (HPV), Hydrogen Peroxide gas plasma (HPGP), Ethylene Oxide (EtO), Liquid Hydrogen Peroxide (LHP), Microwave Oven Irradiation (MOI), Microwave Oven generated Steam (MGS), Moist Heat Incubation (MHI, pasteurization), and Sodium Hypochlorite (bleach, 0.6%) ^2,3^. A study by Visusi et al., used one-cycle (1x) to investigate five decontamination methods [UVGI, HPGP, EtO, MOI, and Sodium Hypochlorite (bleach)] for nine models of NIOSH-certified FFRs, and suggested that UVGI, HPV, and EtO were the most promising methods ^2^. Although these three options emerged as their top choices, none of the five methods negatively affected the structural integrity of the FFRs. Investigations by Bergman et al., examined three-cycle (3x) processing for eight different decontamination methods [UVGI, HPV, HPGP, EtO, LHP, MOGS, MHI, and sodium hypochlorite (bleach, 0.6%)] for six models of FFRs ^3^. They discovered that all the methods for all six FFRs maintained the optimal levels of filter aerosol penetration (<5%), excect for HPGP which had >5% penetration levels for four of the six FFRs. Neither of the two studies, however, examined organism killing as part of the experimental paradigm.

One published report from an FDA award to Battelle Memorial Institute investigated decontamination of N95 FFRs (3M model 1860) using hydrogen peroxide vapor (up to 50 cycles) delivered from a Bioquell Clarus C HPV decontamination system ^4^. The study found that aerosol collection efficiency and air flow resistance were not affected over the 50 cycles of reprocessing. Although no visible degradation of the elastic straps was observed at up to 20 cycles, after 30 cycles the elastic straps showed signs of fragmentation upon stretching. The Battelle study also measured decontamination properties using a biological indicator (BI), *Geobacillus stearothermophilus*, since this spore-forming organism has resistance to HPV decontamination and heat, and therefore, represents a high stringency surrogate for pathogen inactivation. Importantly, their work demonstrated biological aerosol exposure and HPV decontamination were effective for up to 50 cycles with a 6-log reduction in the BI. Battelle recently received approval by the FDA to incorporate the VHP method into a mobile Critical Care Decontamination System™ (CCDS) for large-scale decontamination of PPE for reuse, including N95 respirators for up to 20 cycles ^5^. In line with the Battelle findings, Duke University and Health System recently evaluated and implemented VHP methods for the decontamination and reuse of N95 respirators for up to 30 cycles ^6^. The University of Nebraska Medical Center recently developed a detailed workflow for decontamination of N95 respirators and opted to utilize a UVGI process ^7^. Deployment of reprocessed FFRs for some of their HCP has already been implemented.

Based on the available literature and consultation with subject matter experts throughout the planning phase, we prioritized VHP decontamination of FFRs as a top-choice by mid-February, and subsequently began developing our processes. Additional influence for our choice included: 1) HPV technology is a widely used industry standard for decontamination/sterilization in research and medical facilities, and 2) improved hydrogen peroxide has the lowest EPA acute toxicity category (i.e., category IV) meaning that it is essentially non-toxic and not an irritant for oral, inhalation, and dermal routes of administration ^8,9^. For additional validation of our choice for HPV decontamination, the CDC recently released information about FFR decontamination and reuse as a “crisis capacity strategy to ensure continued availability”, and HPV was listed as one of the most promising potential methods ^10^.

### Collection and Storage of used FFRs

We employed a process in which the HCP removes the FFR following the appropriate institutional guidelines. Inspection for visible soiling, saturation, or loss of structural integrity is performed, and FFRs that are structurally intact and not visibly soiled or saturated are placed in a designated foot-pedal receptacle containing a biohazard bag. Those FFRs that do not meet the inspection standards are discarded in a separate receptacle using standard institutional procedures. This process is followed by safely doffing of the gloves and hand-hygiene.

Designated personnel retrieve the biohazard bags from the unit when the receptacles become half-full per communication (telephone call) from the originating unit. Information communicated from the unit to the designated pick-up individual includes: unit name, location of bins (e.g., room numbers), and assigned contact person on the unit. The individual retrieving the material follows the designated institutional guidelines for ensuring safety. The biohazard bag being retrieved is placed in another biohazard bag and closed using a zip tie. A sticker is placed on the outside of the bag designating date and unit of origin, followed by transport of material to a locked storage area.

### Removing stored FFRs for Processing

The removal of FFRs from their storage container is performed in the same room where the HPV decontamination process occurs. This step is performed by personnel following institutional PPE donning procedures. Based on the exposure risk, appropriate PPE training (or re-training) is provided to the personnel. When processing the FFRs, personnel wear an N95 respirator, eye protection (googles and face shield), protective disposable clothing that covers their skin and hair, and two pairs of gloves. If a powered air purifying respirator (PAPR) is available, this can be implemented instead of the FFR and eye/face coverings for the personnel performing the procedures. The personnel remove the biohazard bags from their transport container. The zip tie on the bag is cut (bags are not to be cut or torn open) and the contents are gently (and slowly) placed on the processing table. Personnel should not reach into the biohazard bag to retrieve (touch) any items nor should the contents be quickly “dumped” onto the processing table to avoid generating aerosolized particles. FFRs are not be touched by the personnel and large forceps (grippers) should be used to retrieve any FFRs that remain in the biohazard bag. During this process, there is an additional inspection of the FFR so that any visibly soiled (e.g., make-up, lotion, dirt, or biological material) respirators do not undergo the decontamination process and are disposed in a biowaste container. Upon emptying all the contents, the biohazard bag is rolled in upon itself and placed into a biowaste container.

### Placing FFRs on Processing Racks

We have employed 5-6 processing racks (2’ x 7’ with four shelves) for each decontamination process. For placement of the respirators, the processing rack (rolling cart) is placed adjacent to the processing table. The FFRs are hung from one of the elastic straps to the end of an “S” hook with the other of the hook adjoined to the wire shelfing in the process rack (Figure 1). Loading of the FFRs begins from the bottom of the processing rack and proceeds sequentially upwards until reaching the top. Large forceps (grippers) are used to hang the FFRs on the “S” hooks without touching with the gloved hands at any time. The FFRs should not be touching one another and should be spaced far enough apart to allow hydrogen peroxide vapor to surround and permeate all surfaces of the respirator (Figure 1). Once loading of the processing rack is complete, it is then transported to a designated location in the room. This process is repeated until all the racks are loaded with the FFRs. There are approximately 165 FFRs placed on each rack for a throughput of 825-990 N95s (5-6 racks) per processing run (Figure 1). The FFRs currently being processed are 3 M™ 1860 and 1870 N95s, as well as protective eyewear.

**Figure 1.**
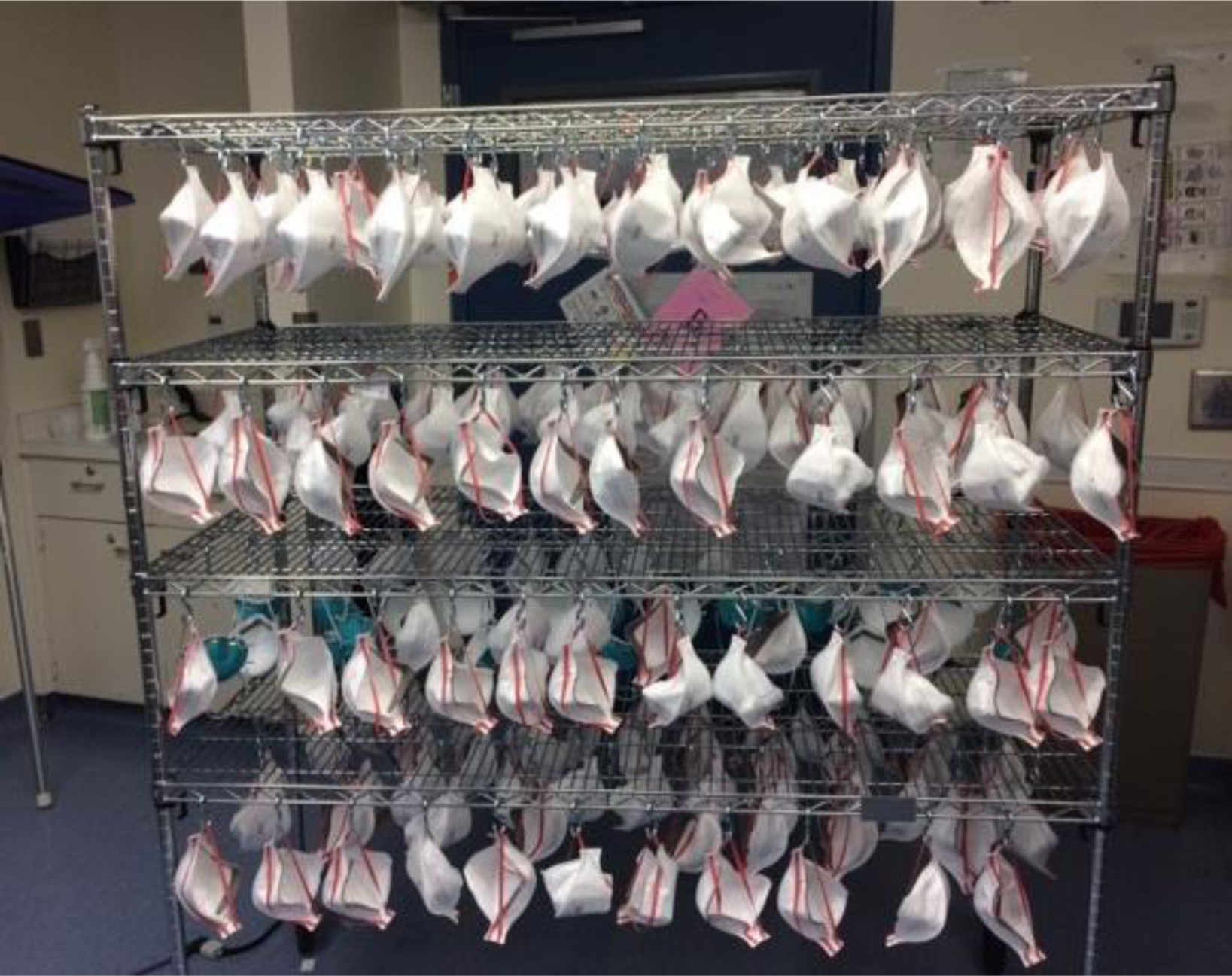
FFR placement and spacing on processing rack.

### HPV Decontamination Process

The HPV sterilization process has been set in an operating room 24’ x 18” with 8’’ ceilings (321 m^3^) (Figure 2). The instrument employed in the room is a Bioquell Clarus™ C hydrogen peroxide vapor generator using 30% w/w H_2_O_2_ solution. The cycle parameters are set with an H_2_O_2_ injection rate and H_2_O_2_ dwell rate for both at 8.0 g/min. The HPV generator has the following phases: Conditioning (10 min), Pre-Gassing, Gassing (83 min), Gassing Dwell (36 min), and Aeration. To facilitate dispersion of the H_2_O_2_ vapor, two circulating fans are placed on the floor of the room (low speed setting) (Figure 2). Each process run in the room includes 4 Bioquel chemical indicators (CIs, Bioquell; Horsham PS), and 10 biological indicators (BIs), *Geobacillus stearothermophilus* spores (Mesa Laboratory; Lakewood, CO), with another BI placed immediately outside of the room to serve as a control. Once the aeration phase is complete, a PortaSens III Hydrogen Peroxide Sensor is used to ensure that H_2_O_2_ vapor in the room is below 1.0 ppm prior to personnel entry into the room ^11^. The CIs were visually inspected immediately after the run and the BIs placed in culture following manufactures instructions. Each run using the conditions listed above has achieved 6-log reduction for the CIs and negative cultures for the BIs (Figure 3). FFRs are not removed from the racks until they reach 0.0 ppm.

**Figure 2.**
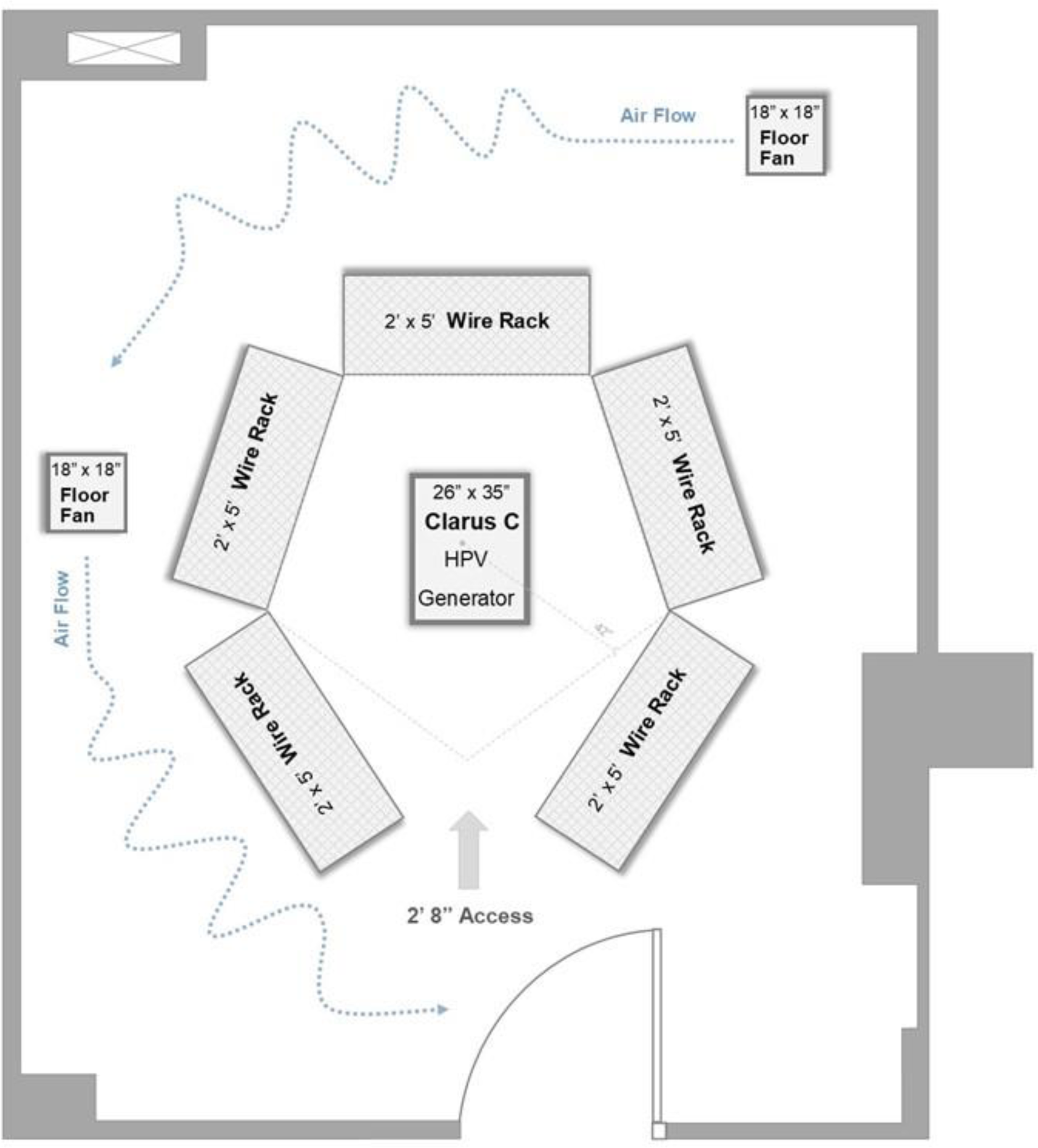
Configuration for HPV decontamination process in an operating room.

**Figure 3.**
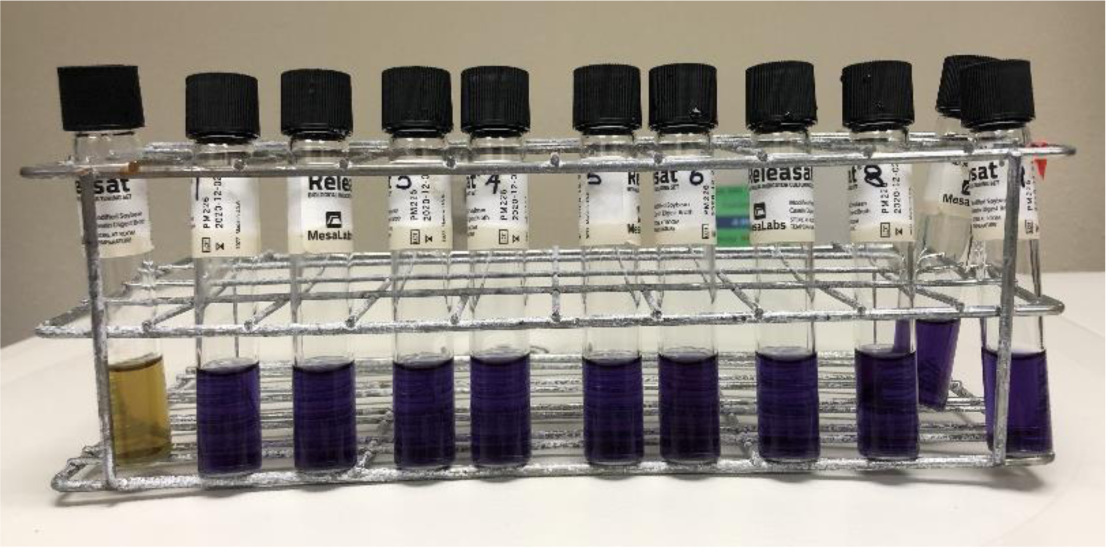
Culture results from biological indicators (*Geobacillus stearothermophilus* spores) with control placed outside the room (left, yellow) and 10 BIs placed in the processing room during the HPV decontamination (right 1-10, purple).

### Post-Processing

The personnel performing the post-processing wear a procedure mask and gloves. Once the FFRs are removed from the rack, they are visibly inspected for any damage, and those with signs of physical damage (mask surfaces, staples, and elastic bands) are discarded. FFRs that pass the physical inspection are marked with a small indelible mark (using a sharpie pen). The marking pattern on the FFRs for up to 20 cycles, the maximum number of reprocessing runs, is shown in Figure 4. The reprocessed FFRs are then placed into individual bags marked with the processing date and batch run, followed by sorting into size and model for redistribution. All users of the reprocessed FFRs should perform a visual inspection of the N95 prior to donning to ensure overall structural integrity, followed by a fit test to ensure that an effective seal is achieved. Those FFRs that do not meet this integrity check are discarded.

**Figure 4.**
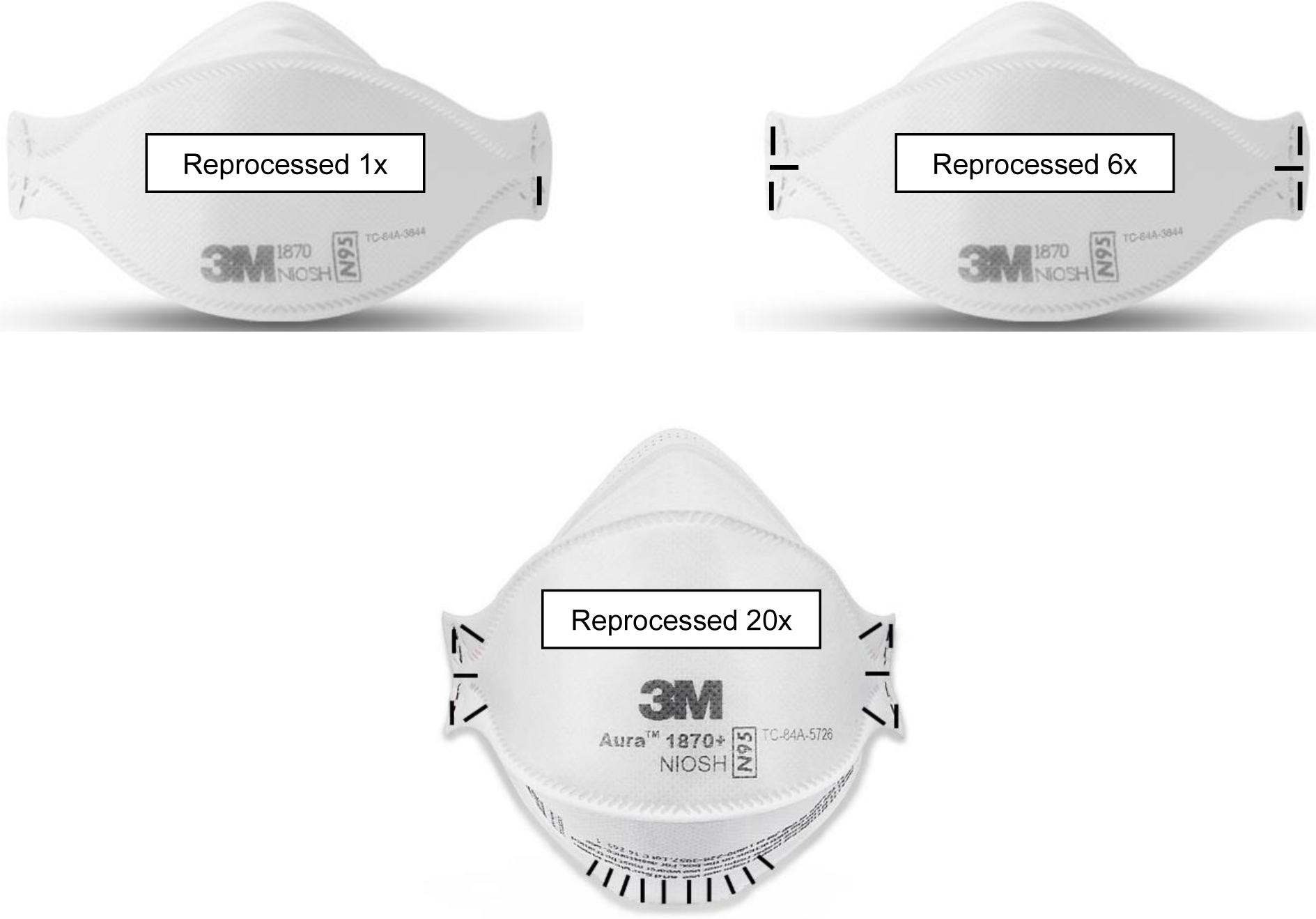
Marking of FFRs to indicate reprocessing cycles.

## CONCLUSION AND DISCUSSION

A short time ago, the decontamination of FFRs for reuse would have been considered (by most) to be either unnecessary or non-viable. However, strain on the global supply chain of PPE, in the context of providing a safe working environment for HCPs, has fostered creative solutions that are now being considered and implemented at some institutions. The most critical steps in the process are: 1) to consider PPE as a limited commodity with a finite supply, and 2) to begin the safe collection and storage of PPE for potential reuse. Without a reserve of supplies to reprocess, the ability to efficiently create a workflow for decontamination and deployment of reprocessed FFRs (or other PPE) becomes exceedingly limited. Prior to deciding on the exact method for the future decontamination procedure that we may needed to implement, we created the workflow to safely collect and store the FFRs (and other PPE) to create sufficient reserves. This allowed us to focus our efforts on deciding which procedure(s) were viable in our environment, and once determined, the ability to rapidly implement the steps involved in the decontamination process.

Based on the available information at the time, we prioritized HPV decontamination as our first choice, and UVGI as a viable second option. However, since we did not have any pre-existing configurations that contained large chambers with external sources of HPV, we started testing in HPV generators in different environments. Learning through trial and error, in an iterative process and with open minds, was critical to our eventual success. Initially, we tested the process in a standard room (22’ x 8’ with 8’ ceilings; 131 m^3^) and were meet with challenges. For example, the room did not have adequate airflow to cool the environment to an optimal temperature between the HPV processing runs. This resulted in the Bioquell instrument shutting down during the gassing phase due to overheating, thereby, reducing the desired levels of H_2_O_2_ (ppm). It became apparent that waiting for a protracted period to allow the room to reach the desired temperature for a subsequent run would not achieve desired efficiently. As such, we eliminated this environment as a viable option and set up the HPV decontamination process in one of four unused operating rooms. Based on their intended use, such environments are constructed with optimized climate control, outside air exchanges, and finishes that are monolithic, scrubbable, and free of crevices and fissures. Sterilization of operating rooms with portable HPV generators, such as the instrument we employed, is an industry standard for no-touch disinfection of the environment to prevent transmission of pathogens. During the HVP exposure application we further isolated the operating room by sealing off the heating, ventilation and air conditioning (HVAC) supply / exhaust ducts and door with polyethylene sheeting and tape. Upon setting up the HPV process in the operating room, we achieved immediate success and moved forward in that setting. We have achieved similar efficacy in a second operating room with a different Bioquell system (BQ-50), indicating flexibility in the overall process.

Results presented in this manuscript are meant to serve as an information sharing tool for other institutions who may wish to set up such processes, particularly for those who do not already have specific HPV chambers already in place. The workflow described here is one of many different options to operationalize the overall process. It is realized that different institutions will have creative ways to find solutions for their own unique challenges with PPE shortages. The two most important lessons learned from our experience are: 1) develop an adequate reserve of PPE for efficiently implementing the reprocessing workflow, and 2) locate a suitable environment for the HPV decontamination procedure, such as an operating room, which has the pre-existing conditions required for conducing the HPV decontamination process. While it is certain that we face unique challenges with COVID-19 that were not previously imagined, an efficient and safe workflow for reprocessing FFRs, and other PPE, can foster substantial improvements for protecting our HCP during this phase of critical shortages. An efficient and robust reprocessing workflow can also promote re-implementation of previous (more stringent) standards of PPE use that were commonly used before the current shortage.

## Data Availability

All data will be made available.

## ACKNOWLEDGEMENTS

We sincerely thank the following individuals at the University of New Mexico Hospital who made suggestions and contributions to protocol development: Jessie D. Pinello, CIC, Infection Preventionist; Amanda V. Martinez, BSN, RN-BC, Infection Preventionist; Shamima Sharmin, MBBS, MSc, MPH, CIC; Meghan Brett, MD, MPH, UNMH Hospital Epidemiologist; Juan Flores, Director UNMH Environmental Services; Anita Nevarez, MBA, Manger UNMH Environmental Services; Aldo Davila, Supervisor UNMH Environmental Services; and Jeremy Jerge, RA I AIA I CCCA. We are also grateful to Tammy Fogg, RN, MSc Nursing, MBA, Executive Consultant for Interim Leadership, B.E. Smith^®^, who oversees the HPV processes; and the certified Bioquell Technicians, Gene Anzures, Joseph Peters, Brian Childs, who perform the reprocessing procedures. The team is also grateful for the support and encouragement by Dr. Richard Larson, MD, PhD, Executive Vice Chancellor, UNMHSC; and Dr. Mark Unruh, MD, MS, Chair of Department of Internal Medicine, UNMHSC.

